# From Mutation to Prognosis: AI-HOPE-PI3K Enables Artificial Intelligence-Agent Driven Integration of PI3K Pathway Data in Colorectal Cancer Precision Medicine

**DOI:** 10.1101/2025.05.25.25328304

**Authors:** Ei-Wen Yang, Brigette Waldrup, Enrique Velazquez-Villarreal

## Abstract

**Introduction:** The incidence of early-onset colorectal cancer (EOCRC) is rising rapidly, with disproportionate health burdens falling on populations that experience both the steepest increases and the poorest outcomes. The phosphoinositide 3-kinase (PI3K) signaling pathway is a key oncogenic driver in colorectal cancer (CRC), influencing tumor growth, survival, and therapeutic resistance. Despite its biological significance, the role of PI3K pathway alterations in EOCRC remains poorly understood—particularly in underrepresented populations—due to limited diversity in genomic datasets and a lack of tools for integrative, pathway-specific analysis. To address this gap, we developed AI-HOPE-PI3K, a conversational artificial intelligence (AI) system designed to streamline clinical-genomic integration and enable real-time, population-aware analysis of PI3K dysregulation in CRC.

**Methods:** AI-HOPE-PI3K is built on a fine-tuned biomedical LLaMA 3 large language model (LLM) and supports natural language queries that are translated into executable statistical pipelines. The platform harmonizes and integrates data from cBioPortal, encompassing key clinical features such as age, race/ethnicity, MSI status, tumor site, stage, treatment history, and survival outcomes. It automates cohort construction, survival modeling, mutation frequency comparison, and odds ratio analysis, delivering interpretive visual and tabular outputs. Validation was performed through the replication of known PI3K-related associations and comparative benchmarking against existing bioinformatics platforms.

**Results:** AI-HOPE-PI3K enabled real-time interrogation of CRC datasets, producing interpretable results without the need for programming. Among EOCRC patients, the frequency of PI3K alterations was similar across different population cohorts. However, colon tumors harboring PI3K alterations were associated with significantly worse survival compared to rectal tumors (p = 0.0177). High tumor mutational burden (TMB) predicted improved survival in FOLFIRI-treated CRC patients (p = 0.0032) and was enriched for MTOR mutations. Among MSI-high patients receiving pembrolizumab, survival did not differ significantly by PIK3CA mutation status. Notably, INPP4B mutations were significantly enriched in H/L EOCRC patients (OR = 3.57, p = 0.005), indicating a potential ancestry-linked biomarker. Analyses stratified by age and stage in PTEN- and PI3K-altered CRC cohorts revealed context-dependent trends in survival.

**Conclusions:** AI-HOPE-PI3K is a first-in-class, conversational AI platform that enables natural language–based, PI3K-pathway-specific analysis of CRC genomics. By integrating multi-institutional datasets with clinical annotations, it democratizes access to complex analyses and enables equitable exploration of PI3K biology. The system reliably reproduced established findings and uncovered novel, population-specific genomic insights—particularly among H/L EOCRC patients. AI-HOPE-PI3K demonstrates the power of AI-driven platforms to advance precision oncology and address disproportionate health burdens through scalable, real-time, and hypothesis-driven clinical-genomic investigation.

## Introduction

Rising rates of early-onset colorectal cancer (EOCRC) have altered the epidemiological landscape of gastrointestinal malignancies. [1–4] Once predominantly a disease of older adults, CRC is increasingly affecting individuals under age 50, with disproportionate impact on specific populations in the United States [5–8]. These trends are not only concerning—they are biologically and clinically distinct [9–11]. EOCRC often presents with unique molecular features and at more advanced stages, suggesting missed opportunities for early detection and a critical need for improved precision oncology strategies [12–15].

Among the most frequently dysregulated molecular cascades in colorectal cancer is the phosphoinositide 3-kinase (PI3K) signaling pathway. A master regulator of growth, survival, and cellular metabolism, PI3K is commonly altered in CRC via mutations in PIK3CA, loss of PTEN, amplification of IGF2, and mutations in AKT1, contributing to tumor progression, therapeutic resistance, and poor outcomes [16–22]. Notably, PI3K dysregulation is implicated in resistance to anti-EGFR therapies and in downstream activation of mTORC1—a driver of metabolic reprogramming and chemoresistance [17,23–26].

Despite its clinical relevance, the full impact of PI3K pathway alterations in EOCRC— particularly among H/L patients—remains underexplored. This knowledge gap stems in part from limited inclusion of diverse populations in large genomic datasets and from a lack of tools that can synthesize clinical, genomic, and demographic data in a targeted, pathway-specific manner.

Conventional platforms such as cBioPortal [27] and UCSC Xena [28] offer robust data access but fall short in three key areas: (1) real-time hypothesis testing, (2) user-friendly interface for non-programmers, and (3) contextualized analysis that incorporates variables such as ethnicity [29–30], MSI status [31–33], and treatment history [34–38]. These limitations hinder precision medicine efforts aimed at addressing molecular drivers of disease in underserved populations.

Recent breakthroughs in artificial intelligence (AI)—specifically natural language–driven large language models (LLMs)—offer a new paradigm for clinical-genomic integration [39–47]. These models are capable of interpreting user queries and converting them into executable analytical pipelines, democratizing data access and enabling hypothesis generation without the need for coding expertise.

To harness this potential for cancer research, we developed AI-HOPE-PI3K—a conversational AI system designed to investigate PI3K pathway alterations in CRC using harmonized clinical and genomic data. Unlike generic analytic platforms, AI-HOPE-PI3K was engineered to perform pathway-centered analyses in response to natural language prompts, stratify patient cohorts by age, ancestry, and molecular profiles, and automate survival and association testing across large datasets.

In this study, we present the development and deployment of AI-HOPE-PI3K, validate its performance through reproduction of known PI3K associations [1], and apply it to uncover novel patterns in EOCRC among patients from different population cohorts. Our findings underscore the value of AI-driven, pathway-specific tools in accelerating translational cancer research and addressing disparities in molecular oncology.

## Methods

### Overview of AI-HOPE-PI3K Design Philosophy

AI-HOPE-PI3K was developed to bridge the gap between advanced computational analysis and the practical needs of translational cancer researchers. It is a user-facing, natural language–driven platform specifically designed to support integrative, pathway-centric investigations of PI3K signaling dysregulation in CRC. Central to its architecture is a conversational interface that translates biological questions into rigorous, interpretable, and reproducible analyses—without requiring programming expertise or manual scripting (Figure 1).

**Figure 1.**
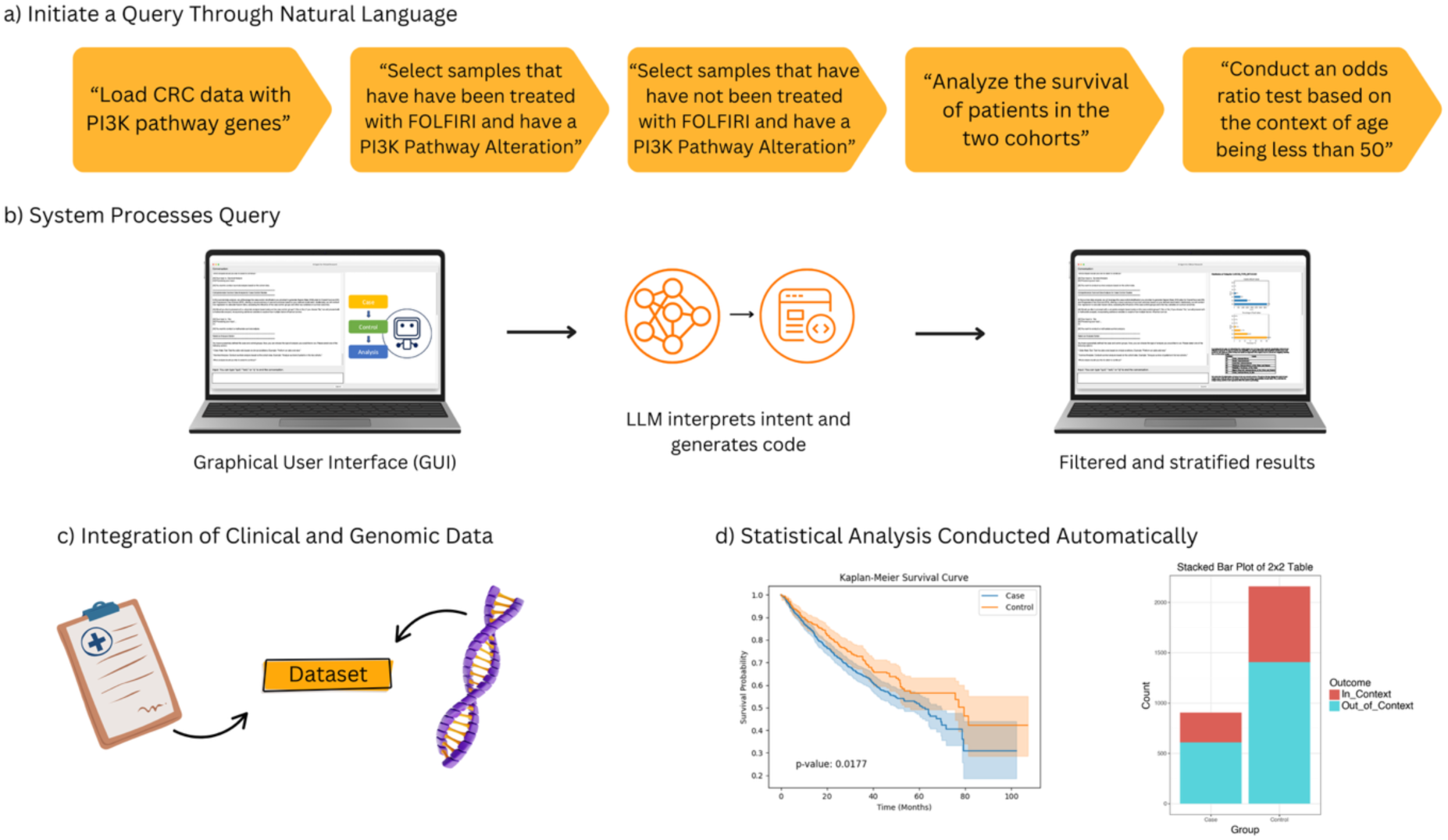
Overview of the AI-HOPE-PI3K Workflow for Pathway-Specific Precision Oncology in Colorectal Cancer (CRC). This figure illustrates the end-to-end architecture and functionality of AI-HOPE-PI3K, a conversational artificial intelligence system designed for integrative analysis of PI3K pathway alterations in CRC. (a) Users initiate queries through natural language prompts, such as filtering by treatment exposure (e.g., FOLFIRI), mutation status in PI3K pathway genes (e.g., *PIK3CA*, *PTEN*), and age-related stratification (e.g., patients under 50 years). (b) The system processes the query through a graphical user interface (GUI) that leverages a large language model (LLM) to interpret semantic intent and translate it into executable code, producing filtered and stratified outputs. (c) Harmonized clinical and genomic data are integrated in real time, allowing the system to dynamically generate case-control cohorts for downstream analyses. (d) Statistical analyses, including Kaplan-Meier survival curves and odds ratio tests from contingency tables, are automatically conducted and visualized.

### Natural Language Engine and Semantic Workflow Translation

At the core of AI-HOPE-PI3K lies a fine-tuned biomedical large language model (LLM) built on the LLaMA 3 architecture. When a user poses a question—e.g., “What is the survival impact of PTEN loss in EOCRC among Hispanic/Latino patients?”—the LLM identifies the key intent (e.g., survival, mutation, population), classifies the relevant pathway components (e.g., PTEN, PI3K/AKT/mTOR), and generates the corresponding code to execute downstream analysis. A semantic parser then maps variables like age, MSI status, tumor location, and treatment exposure to internal ontology tags. These elements are used to construct dynamically filtered cohorts, which feed into a statistical pipeline for hypothesis testing.

### Data Backbone and Pathway-Centric Structuring

AI-HOPE-PI3K integrates harmonized datasets from TCGA, AACR GENIE, and cBioPortal, preprocessed for compatibility with automated cohort stratification. It focuses on genomic alterations within key PI3K pathway components: PIK3CA, PTEN, AKT1, IGF2, TSC1/2, MTOR, RHEB, and RPTOR. These data are cross-linked with clinical attributes including patient age, race/ethnicity, tumor location (primary/metastatic), stage, MSI status, therapeutic history (e.g., anti-EGFR exposure), and survival duration. Preprocessing involved several steps: Conversion into tabular, analysis-ready matrices Harmonization of patient/sample IDs, Application of ontology labels (e.g., OncoTree disease codes, race/ethnicity categories), Verification of mutation types (e.g., missense vs. truncating) and allele frequencies. This structured backend allows AI-HOPE-PI3K to rapidly generate intersectional cohort definitions for tailored PI3K analyses.

### Statistical Capabilities and Analytical Modules

The platform’s statistical engine was implemented in Python and supports both exploratory and confirmatory analysis workflows. Key functions include: 1) Mutation analysis: Frequency comparisons by subgroup using chi-square or Fisher’s exact tests; 2) Association metrics: Odds ratio estimation with 95% confidence intervals; 3) Survival modeling: Kaplan-Meier estimation with log-rank testin multivariable Cox proportional hazards regression where appropriate; 4) Stratified analysis: Automated subgrouping based on combinations of mutation status, MSI, race/ethnicity, tumor site, and age group; 5) Molecular patterning: Co-occurrence/mutual exclusivity testing among PI3K genes and pathway cross-talk with other molecular characteristics. Each query triggers the generation of plots, tables, and natural language summaries that interpret the results within a clinical-translational context.

### Validation and Use Case Scenarios

To validate AI-HOPE-PI3K’s analytical accuracy and biological relevance, we reproduced previously established associations involving PIK3CA mutations and poor survival, PTEN loss and resistance to anti-EGFR therapy, and PI3K-driven mTOR activation in chemo resistant CRC [1]. Additional real-world use cases included survival comparisons between H/L and NHW EOCRC patients with AKT1 mutations, PI3K mutation frequency across MSI-high vs. MSS tumors, and odds ratio testing of PTEN deletions stratified by therapy exposure. These validation exercises confirmed the platform’s ability to replicate known findings while supporting exploratory hypothesis generation.

### User Interaction and Performance Benchmarking

AI-HOPE-PI3K was benchmarked against cBioPortal and UCSC Xena using real-world tasks. The evaluation focused on three criteria: (1) ability to interpret complex prompts involving age, race, mutation, and MSI simultaneously; (2) speed of cohort generation and statistical output; and (3) clarity of generated visuals and summaries. AI-HOPE-PI3K demonstrated superior responsiveness and interpretability, particularly in multi-variable, disproportionate health burdens-focused analyses.

### Visualization and Result Dissemination

Outputs from AI-HOPE-PI3K include high-resolution figures and annotated data tables: Kaplan-Meier survival curves with p-values and sample counts, forest plots for odds ratios and confidence intervals, bar charts and co-mutation heatmaps for frequency analysis and summary tables ready for supplemental materials or figure legends. All visualizations are created using Matplotlib, Seaborn, and Plotly. Export options include CSV (for tables), PNG (for figures), and narrative-ready PDF summaries for direct use in manuscripts or presentations.

## Results

AI-HOPE-PI3K enables seamless clinical-genomic interrogation of PI3K pathway dysregulation in CRC by translating natural language prompts into fully automated analyses. Users can generate stratified case-control cohorts and statistical outputs including Kaplan-Meier survival curves, mutation frequency comparisons, and odds ratio calculations without coding expertise. Across validation and exploratory tasks, the platform reproduced known associations and revealed novel patterns, particularly in EOCRC and H/L subgroups.

In ancestry-stratified analyses, AI-HOPE-PI3K evaluated the prevalence of PI3K pathway alterations among EOCRC patients across racial/ethnic backgrounds (Figure 2). Among 153 H/L patients and 1,117 NHW patients under age 50, PI3K pathway alterations were observed in 35.29% of H/L and 33.03% of NHW cases. The odds ratio was 1.106 (95% CI: 0.776–1.576; p = 0.642), indicating no statistically significant difference. These results suggest no differential enrichment of PI3K mutations by ethnicity in this EOCRC cohort.

**Figure 2.**
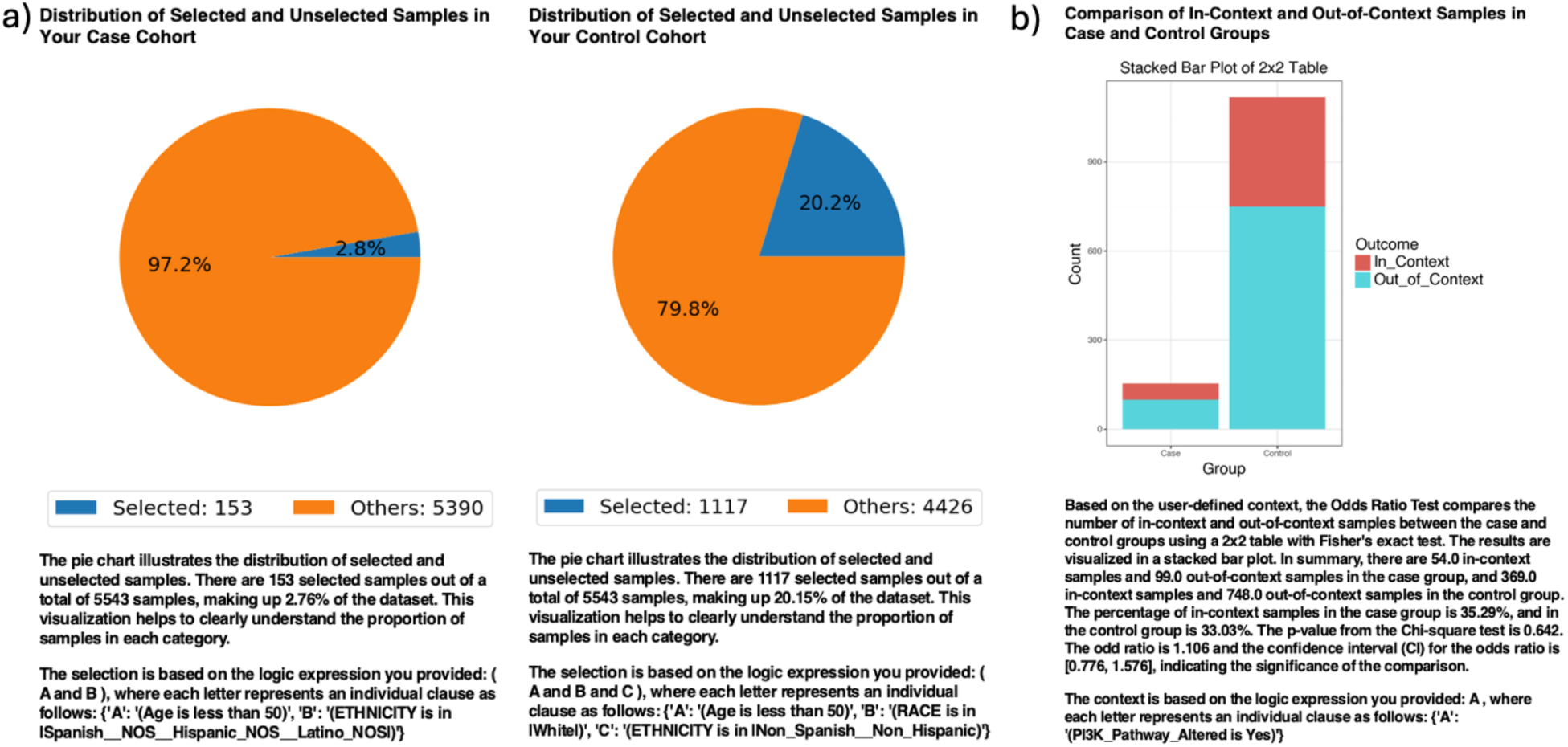
AI-HOPE-PI3K analysis of PI3K pathway alterations in early-onset colorectal cancer (EOCRC) among Hispanic/Latino (H/L) and Non-Hispanic White (NHW) patients. a) Pie charts display the proportion of selected samples in each cohort after natural language–guided filtering of the harmonized dataset. The case cohort includes 153 EOCRC patients under age 50 identified as H/L, representing 2.76% of the dataset. The control cohort includes 1,117 EOCRC patients under age 50 identified as Non-Hispanic White, representing 20.15% of the dataset. b) A 2×2 odds ratio analysis evaluates the frequency of PI3K pathway alterations between the two groups. The stacked bar plot illustrates the distribution of samples with and without PI3K pathway alterations, labeled as “In_Context” and “Out_of_Context,” respectively. PI3K pathway alterations were present in 35.29% of H/L cases and 33.03% of NHW controls. The calculated odds ratio was 1.106 (95% CI: 0.776–1.576), with a p-value of 0.642, indicating no statistically significant difference.

Exploring anatomical differences, AI-HOPE-PI3K stratified CRC cases by primary tumor site—colon versus rectum—among patients harboring PI3K alterations (Figure 3). The case cohort included 977 colon tumor cases; the control cohort comprised 343 rectal tumor cases. Kaplan-Meier survival analysis demonstrated significantly worse outcomes in the colon subgroup (p = 0.0177). This finding highlights a potential prognostic role of tumor location among PI3K-mutated CRC patients.

**Figure 3.**
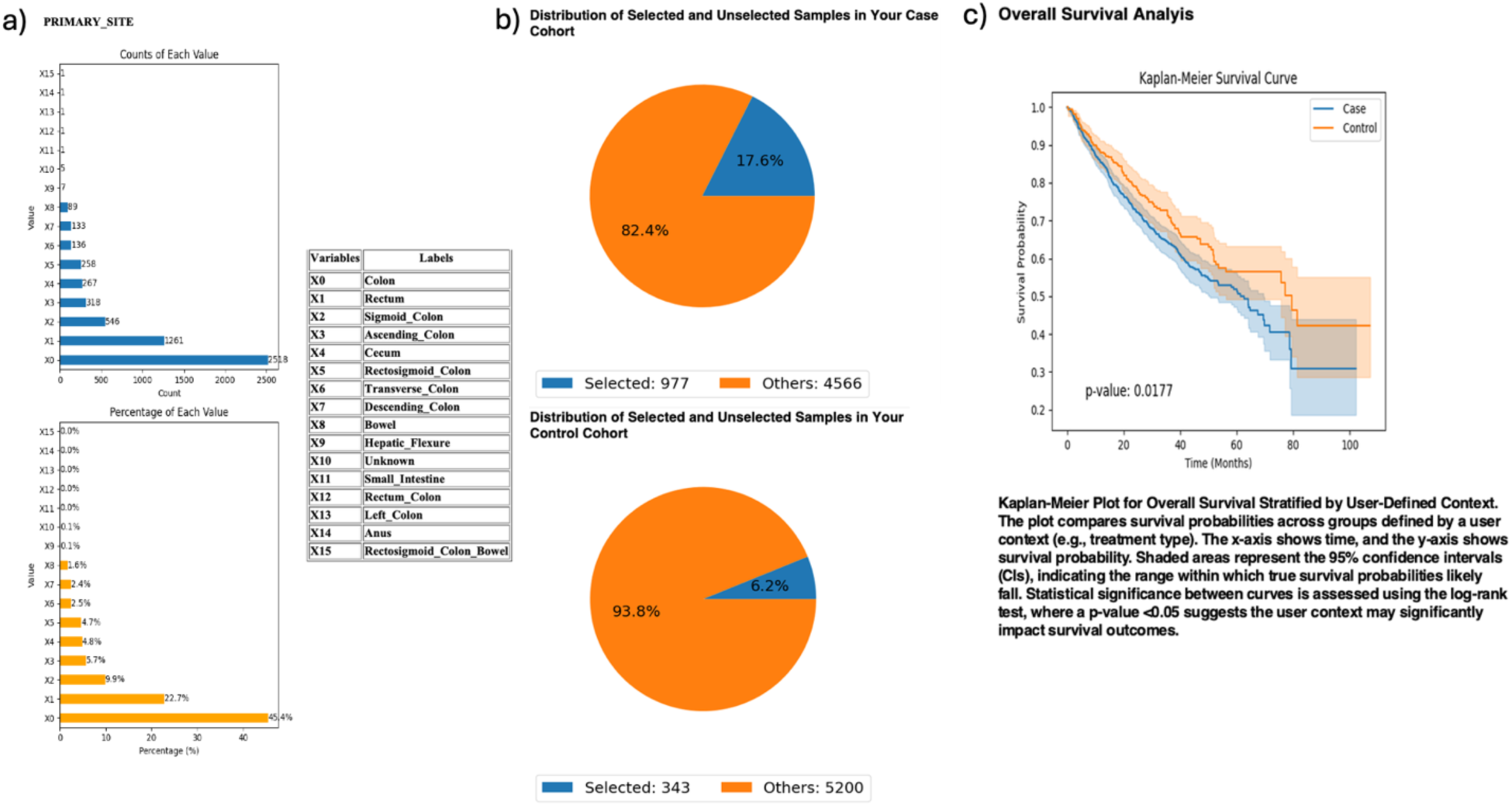
AI-HOPE-PI3K analysis of PI3K-altered colorectal cancer (CRC) samples by primary tumor location (Colon vs. Rectum). This figure illustrates the application of AI-HOPE-PI3K to evaluate survival outcomes among CRC patients with PI3K pathway alterations, stratified by primary tumor site. a) Bar plots display the distribution of tumor site annotations across the dataset, with “Colon” (X0) and “Rectum” (X1) representing the most frequent anatomical sites. Colon samples constitute the majority of annotated entries (45.4%), while rectal tumors account for 22.7%. All other primary sites are shown for context. b) Pie charts visualize the subset of samples selected through natural language–driven query filters. The case cohort includes 977 CRC patients with PI3K-altered tumors located in the colon (17.6% of the dataset), while the control cohort consists of 343 patients with PI3K-altered tumors in the rectum (6.2% of the dataset). c) Kaplan-Meier survival curves compare overall survival between the two groups. CRC patients with PI3K pathway alterations originating in the colon exhibited significantly worse survival outcomes compared to those with rectal tumors (p = 0.0177). Shaded regions represent 95% confidence intervals.

AI-HOPE-PI3K was further applied to examine tumor mutational burden (TMB) and its relationship with MTOR mutation status and survival in FOLFIRI-treated CRC patients (Figure 4). Patients with high TMB (>10; n = 466) had significantly better survival outcomes than low-TMB counterparts (n = 3,257), with a p-value of 0.0032. Odds ratio testing showed MTOR mutations were more frequent in the high-TMB group, suggesting a biologically relevant association between TMB and MTOR in the context of chemotherapy response.

**Figure 4.**
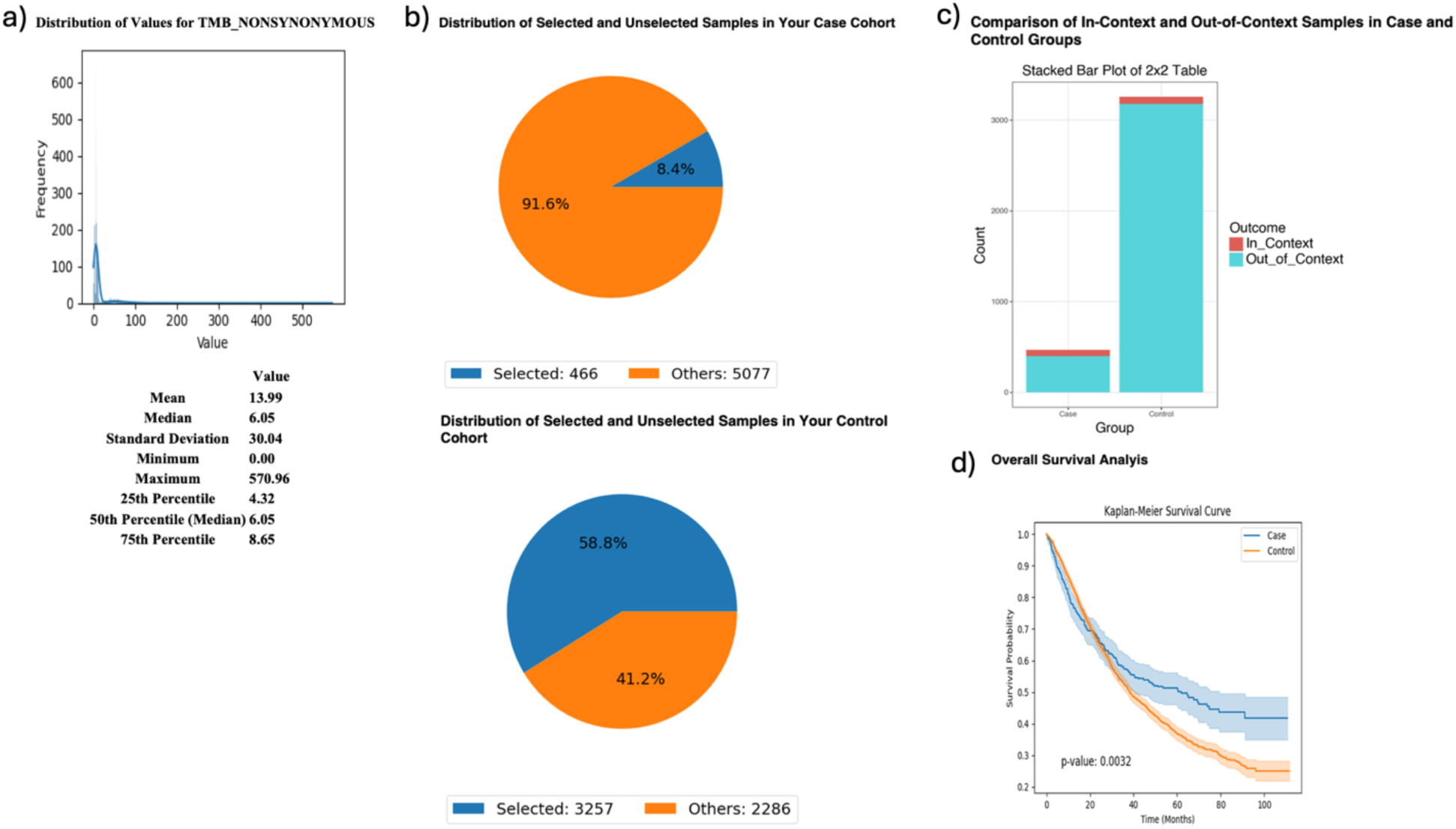
AI-HOPE-PI3K analysis of tumor mutational burden (TMB) and survival outcomes in colorectal cancer (CRC) patients treated with FOLFIRI chemotherapy, stratified by MTOR mutation status. This figure demonstrates the use of AI-HOPE-PI3K to evaluate survival and mutation context in CRC patients treated with the FOLFIRI regimen (Fluorouracil, Leucovorin, Irinotecan), stratified by TMB and analyzed for MTOR mutation enrichment. a) A histogram displays the distribution of nonsynonymous TMB across all samples. The mean TMB is 13.99, with a median of 6.05 and a long right-tailed distribution. These metrics contextualize the threshold (TMB >10) used to define the high-TMB case cohort. b) Pie charts show the distribution of selected samples within each cohort following query-based stratification. The case cohort includes 466 high-TMB CRC patients treated with FOLFIRI (8.4% of the dataset), while the control cohort includes 3,257 low-TMB CRC patients treated with the same regimen (58.8% of the dataset). c) A 2×2 odds ratio analysis evaluates the enrichment of MTOR mutations between the two groups. The stacked bar plot shows that MTOR mutations were more frequently observed in the high-TMB group. However, detailed statistical outputs are not shown here. d) Kaplan-Meier survival curves compare overall survival between high-TMB and low-TMB patients, both treated with FOLFIRI. Patients with high TMB exhibited significantly improved survival outcomes (p = 0.0032). Shaded areas represent 95% confidence intervals, indicating a robust survival benefit associated with elevated TMB in the context of FOLFIRI chemotherapy.

To investigate PI3K immunotherapy response interactions, AI-HOPE-PI3K analyzed MSI-high CRC patients treated with pembrolizumab, comparing those with and without PIK3CA mutations (Figure 5). The case cohort included 52 PIK3CA-mutant samples; the control group had 60 wild-type samples. Survival analysis revealed no significant difference between groups (p = 0.3054), though PIK3CA-mutated patients showed a trend toward improved outcomes.

**Figure 5.**
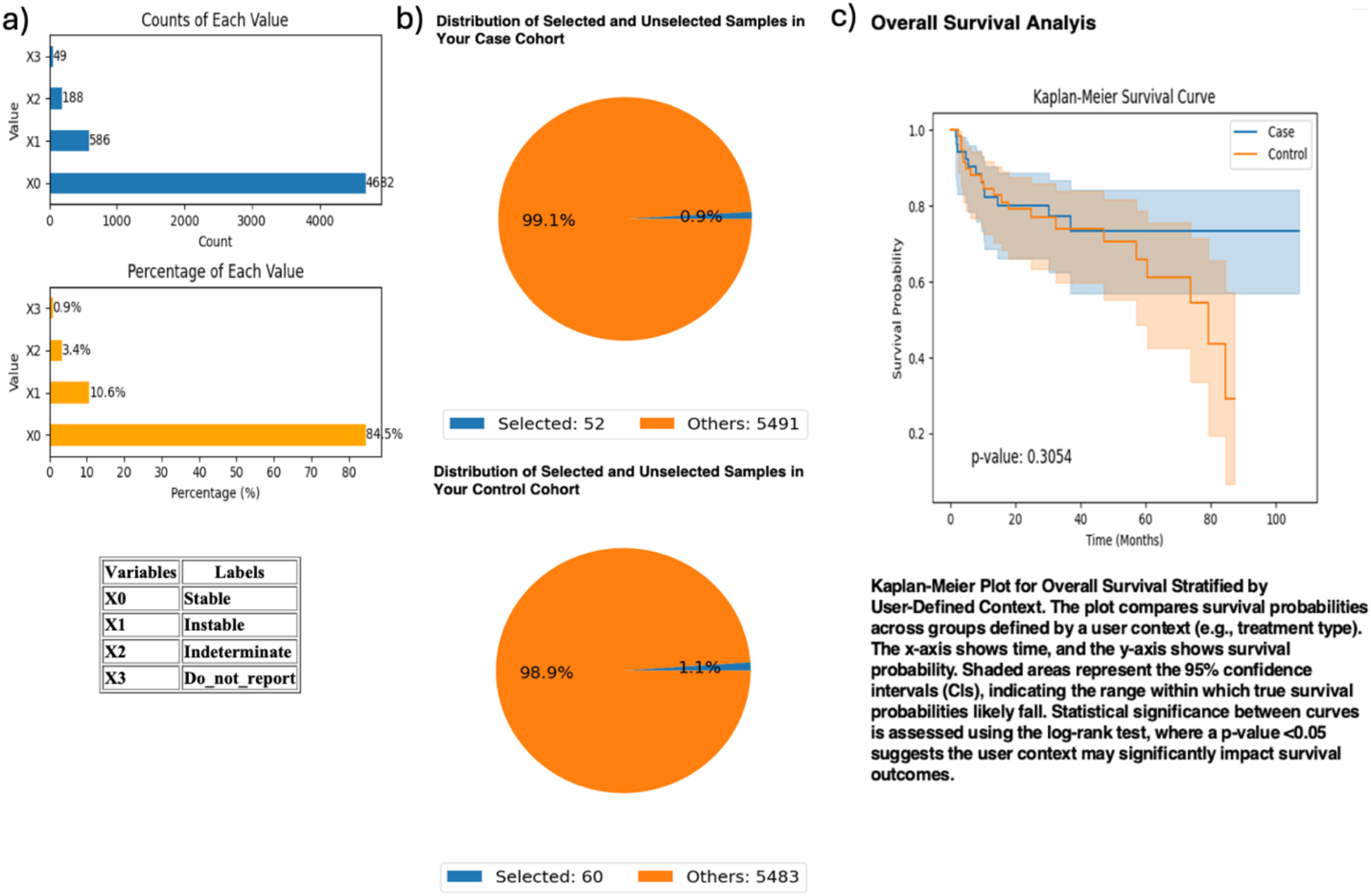
AI-HOPE-PI3K analysis of PIK3CA mutation status among microsatellite instability-high (MSI-H) colorectal cancer (CRC) patients treated with pembrolizumab. This figure demonstrates the use of AI-HOPE-PI3K to assess survival outcomes in MSI-H CRC patients receiving immunotherapy, stratified by PIK3CA mutation status. a) Bar plots show the overall distribution of microsatellite instability types in the dataset. Samples classified as “Instable” (X1) account for 10.6% (n = 586), while “Stable” (X0) represent the majority (84.5%, n = 4,682). Other MSI classifications, including “Indeterminate” and “Do_not_report,” appear less frequently. b) Pie charts visualize cohort selection following natural language query execution. The case cohort includes 52 MSI-H CRC patients with PIK3CA mutations treated with pembrolizumab (0.9% of the dataset), while the control cohort includes 60 MSI-H patients without PIK3CA mutations treated with the same agent (1.1% of the dataset). c) Kaplan-Meier survival analysis compares overall survival between the two groups. While PIK3CA-mutant MSI-H patients appear to have improved survival relative to wild-type counterparts, the difference was not statistically significant (p = 0.3054). Shaded regions represent the 95% confidence intervals, suggesting overlapping survival distributions and insufficient evidence of differential outcomes by PIK3CA status in this immunotherapy-treated subgroup.

Supplementary analyses identified gene-specific disparities among EOCRC subgroups. INPP4B mutations were significantly enriched in H/L versus NHW EOCRC patients (5.23% vs. 1.52%, OR = 3.57, 95% CI: 1.514–8.419, p = 0.005; Figure S1), suggesting a potential ancestry-linked biomarker. While AKT1 (Figure S2) and TSC1 (Figure S3) mutations showed higher rates in H/L cases (OR = 2.19 and 2.00, respectively), neither reached statistical significance.

In age-stratified survival analyses, AI-HOPE-PI3K assessed PTEN-mutated CRC patients treated with FOLFOX chemotherapy (Figure S4). Patients under 50 (n = 59) exhibited a nonsignificant trend toward improved survival compared to those over 50 (n = 122; p = 0.1758), suggesting potential age-modified outcomes in PTEN-mutated subgroups.

Finally, AI-HOPE-PI3K explored stage-specific prognostic variation in CRC patients with PI3K pathway alterations receiving FOLFOX (Figure S5). Comparing early-stage (n = 628) to advanced-stage (n = 507) cases, no significant survival difference was found (p = 0.1267). Odds ratio analysis indicated higher but non-significant enrichment of PI3K alterations in early-stage disease (OR = 1.23, p = 0.174).

Collectively, these results highlight the versatility of AI-HOPE-PI3K in executing diverse, context-aware clinical-genomic analyses. The platform successfully recapitulated known relationships—such as TMB-associated survival benefits and site-specific prognostic variation—while uncovering new ancestry-linked mutation patterns. By enabling realtime, population-aware interrogation of PI3K pathway biology, AI-HOPE-PI3K supports precision oncology efforts aimed at identifying clinically actionable biomarkers and reducing the disproportionate health burdens in colorectal cancer outcomes.

## Discussion

This study presents the development and application of AI-HOPE-PI3K, a novel conversational AI system that enables real-time, natural language–driven analysis of PI3K pathway alterations in CRC. Our findings demonstrate the platform’s ability to replicate established clinical-genomic associations, reveal emerging molecular patterns, and support population-aware hypothesis generation in EOCRC, with particular emphasis on disproportionate health burdens affecting specific populations.

AI-HOPE-PI3K addresses long-standing challenges in cancer genomics research, including the lack of user-friendly tools capable of synthesizing molecular, clinical, and demographic variables in a pathway-specific context. By leveraging a fine-tuned biomedical LLM, the system translates natural language queries into executable workflows, eliminating the need for programming expertise and dramatically reducing the time required for exploratory cohort analysis. Importantly, AI-HOPE-PI3K integrates harmonized datasets with standardized clinical annotations, enabling intersectional stratification across variables such as age, MSI status, race/ethnicity, treatment history, and tumor location—factors often underrepresented in traditional analytical pipelines.

In validation tasks, AI-HOPE-PI3K successfully recapitulated key associations previously reported in the literature. For instance, colon tumor location among PI3K-altered CRC patients was significantly associated with poorer survival compared to rectal tumors (p = 0.0177), corroborating prior studies on anatomical variation in CRC outcomes. Similarly, patients with high tumor mutational burden (TMB >10) treated with FOLFIRI exhibited significantly improved survival (p = 0.0032), supporting the clinical relevance of TMB as a predictive biomarker in immunogenic or chemotherapy-sensitive settings.

Crucially, the platform revealed novel insights into ancestry-linked molecular patterns. While PI3K alteration rates were not significantly different between H/L and NHW EOCRC patients, INPP4B mutations were significantly enriched in H/L individuals (OR = 3.57, p = 0.005), suggesting a potential ancestry-specific biomarker that warrants further investigation. This finding aligns with growing recognition that genomic drivers may vary across racial and ethnic populations and highlights the importance of inclusive datasets and tailored analytical approaches in health disparities research.

Exploratory analyses of AKT1 and TSC1 mutations in H/L EOCRC cohorts revealed higher mutation frequencies, although statistical significance was not reached. These trends nonetheless suggest potential biological relevance, particularly when considered alongside the enrichment of INPP4B and the broader role of the PI3K pathway in therapy resistance and immune evasion. Additional studies with larger and more diverse cohorts will be critical to validate these preliminary observations.

In immunotherapy-treated subgroups, AI-HOPE-PI3K evaluated survival among MSI-H CRC patients receiving pembrolizumab, stratified by PIK3CA mutation status. Although no statistically significant difference was observed, the analysis identified a trend toward improved survival in PIK3CA-mutant cases, consistent with hypotheses suggesting enhanced immunogenicity in tumors with co-occurring PI3K alterations. This underscores the value of AI-driven tools in rapidly generating context-specific hypotheses that can guide prospective biomarker development and clinical trial design.

Age- and stage-stratified analyses further demonstrated AI-HOPE-PI3K’s capacity to uncover subtle survival trends. While PTEN-mutated early-onset patients showed a non-significant trend toward better outcomes than late-onset cases, and PI3K-altered tumors were more frequently observed in early-stage disease, neither result reached statistical significance. These findings emphasize the complexity of CRC progression and the need for nuanced, multivariate modeling to disentangle interactions among genomic, clinical, and demographic variables.

Together, these results highlight AI-HOPE-PI3K’s value as a versatile and scalable platform for translational cancer research. Beyond facilitating hypothesis testing and pathway-specific exploration, the system empowers researchers to perform real-time, population-aware analyses that incorporate critical social determinants of health—an essential step toward equitable precision medicine. As genomic data generation continues to accelerate, tools like AI-HOPE-PI3K will be increasingly vital in making complex datasets accessible, interpretable, and actionable for diverse biomedical audiences.

Future work will expand AI-HOPE-PI3K’s capabilities to enable communication between this AI agent and other agents within the same framework using Model Context Protocol (MCP) Agent-to-Agent (A2A) communication. These agents will be specifically trained to explore additional pathways (e.g., WNT, TGF-β, TP53) or molecular characteristics (e.g., functional genomics, spatial transcriptomics, spatial proteomics), contributing to the development of a universal AI-agent infrastructure. Additional enhancements will include the incorporation of multi-omics data and deployment in clinical decision-support environments. Furthermore, efforts are underway to extend the platform’s reach to community health settings, ensuring that the benefits of precision oncology are distributed across all populations.

## Conclusion

In conclusion, AI-HOPE-PI3K represents a transformative advancement in pathway-centric precision oncology, enabling intuitive, natural language–driven exploration of PI3K alterations in colorectal cancer. By integrating harmonized clinical and genomic datasets with a biomedical LLM, the system facilitates rapid, accessible, and population-aware analyses that uncover both established and novel molecular associations. Its ability to support intersectional stratification and generate hypothesis-driven insights— particularly in patients from populations with disproportionate health burdens— demonstrates its potential to accelerate cancer discovery for all populations. As AI-HOPE-PI3K continues to evolve, its planned expansion into multi-agent communication, multi-omics integration, and clinical decision-support tools underscores its scalability and future relevance. Ultimately, this platform lays the foundation for an interoperable AI-agent infrastructure that democratizes access to complex biomedical data and fosters inclusive, data-informed precision medicine.

## Data Availability

All data used in the present study is publicly available at https://www.cbioportal.org/ and https://genie.cbioportal.org. Additional data can be provided upon reasonable request to the authors.

---

**Figure S1.**
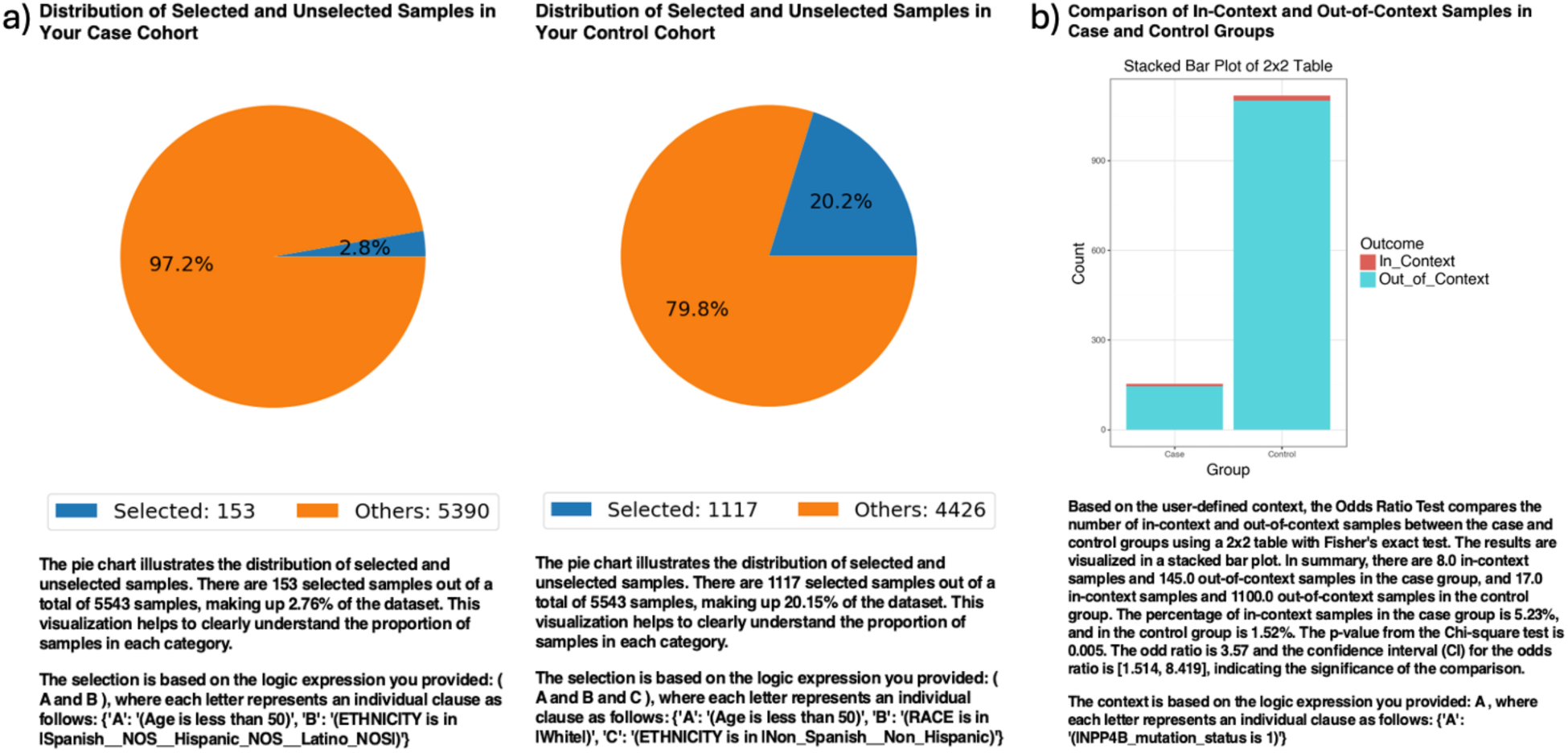
AI-HOPE-PI3K analysis of INPP4B mutations in early-onset colorectal cancer (EOCRC) among Hispanic/Latino (H/L) and Non-Hispanic White (NHW) patients. a) Pie charts illustrate the distribution of selected samples following natural language–driven filtering of the dataset. The case cohort includes 153 EOCRC patients under age 50 with H/L ethnicity, representing 2.76% of the dataset. The control cohort includes 1,117 EOCRC patients under age 50 who are NHW, representing 20.15% of the dataset. b) A 2×2 odds ratio analysis evaluates the frequency of INPP4B mutations between the two groups. The stacked bar plot shows the proportion of samples with and without INPP4B mutations, labeled as “In_Context” and “Out_of_Context,” respectively. INPP4B mutations were present in 5.23% of H/L cases and 1.52% of NHW controls. The calculated odds ratio was 3.57 (95% CI: 1.514–8.419), with a p-value of 0.005, indicating a statistically significant difference.

**Figure S2.**
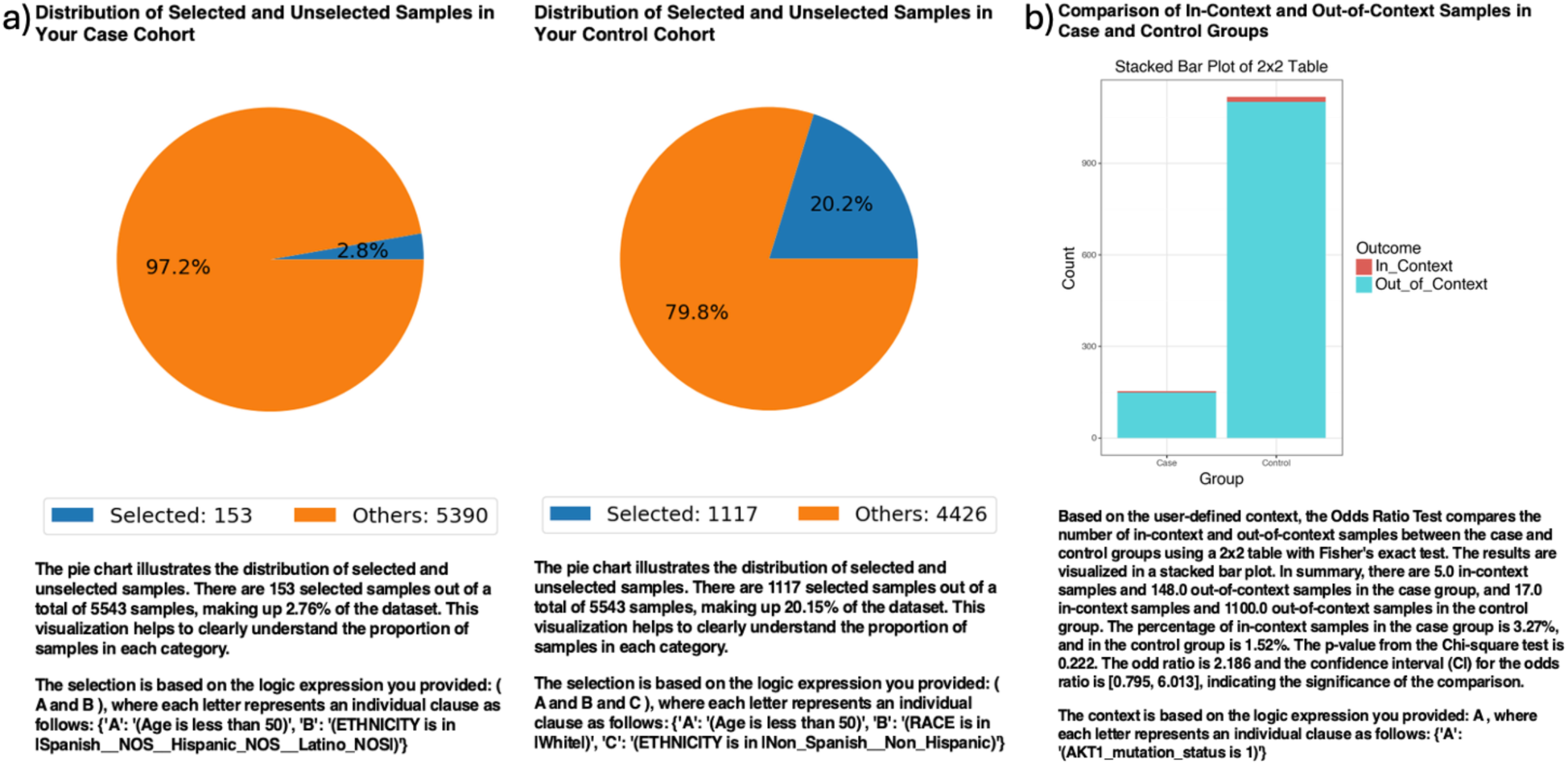
AI-HOPE-PI3K analysis of AKT1 mutations in early-onset colorectal cancer (EOCRC) among Hispanic/Latino (H/L) and Non-Hispanic White (NHW) patients. a) Pie charts illustrate the distribution of selected samples following natural language–driven filtering of the dataset. The case cohort includes 153 EOCRC patients under age 50 with H/L ethnicity, representing 2.76% of the dataset. The control cohort includes 1,117 EOCRC patients under age 50 who are NHW, representing 20.15% of the dataset. b) A 2×2 odds ratio analysis evaluates the frequency of AKT1 mutations between the two groups. The stacked bar plot shows the proportion of samples with and without AKT1 mutations, labeled as “In_Context” and “Out_of_Context,” respectively. AKT1 mutations were present in 3.27% of H/L cases and 1.52% of NHW controls. The calculated odds ratio was 2.19 (95% CI: 0.795–6.013), with a p-value of 0.222, indicating no statistically significant difference.

**Figure S3.**
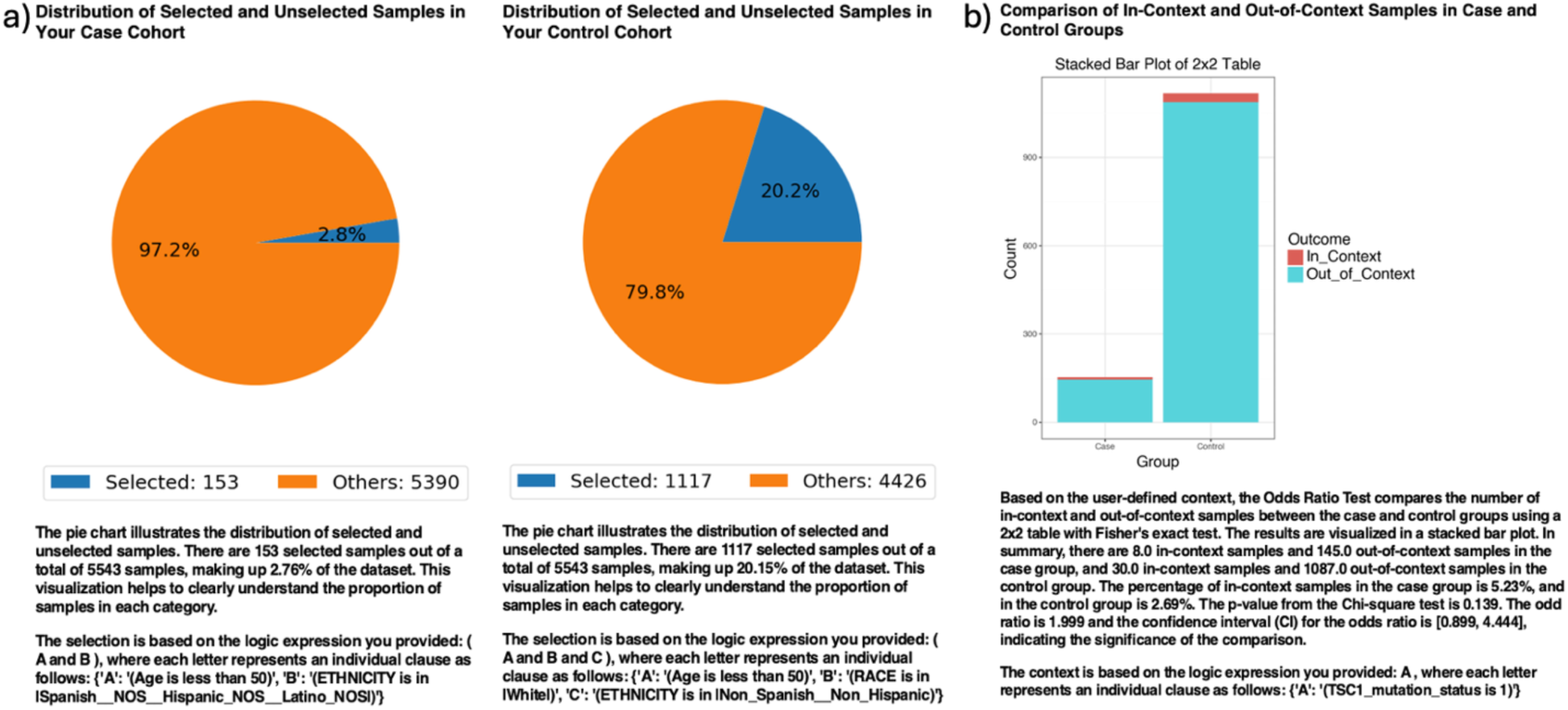
AI-HOPE-PI3K analysis of TSC1 mutations in early-onset colorectal cancer (EOCRC) among Hispanic/Latino (H/L) and Non-Hispanic White (NHW) patients. a) Pie charts illustrate the distribution of selected samples following natural language–driven filtering of the dataset. The case cohort includes 153 EOCRC patients under age 50 with H/L ethnicity, representing 2.76% of the dataset. The control cohort includes 1,117 EOCRC patients under age 50 who are NHW, representing 20.15% of the dataset. b) A 2×2 odds ratio analysis evaluates the frequency of TSC1 mutations between the two groups. The stacked bar plot displays the proportion of samples with and without TSC1 mutations, labeled as “In_Context” and “Out_of_Context,” respectively. TSC1 mutations were present in 5.23% of H/L cases and 2.69% of NHW controls. The calculated odds ratio was 2.00 (95% CI: 0.899–4.444), with a p-value of 0.139, indicating no statistically significant difference.

**Figure S4.**
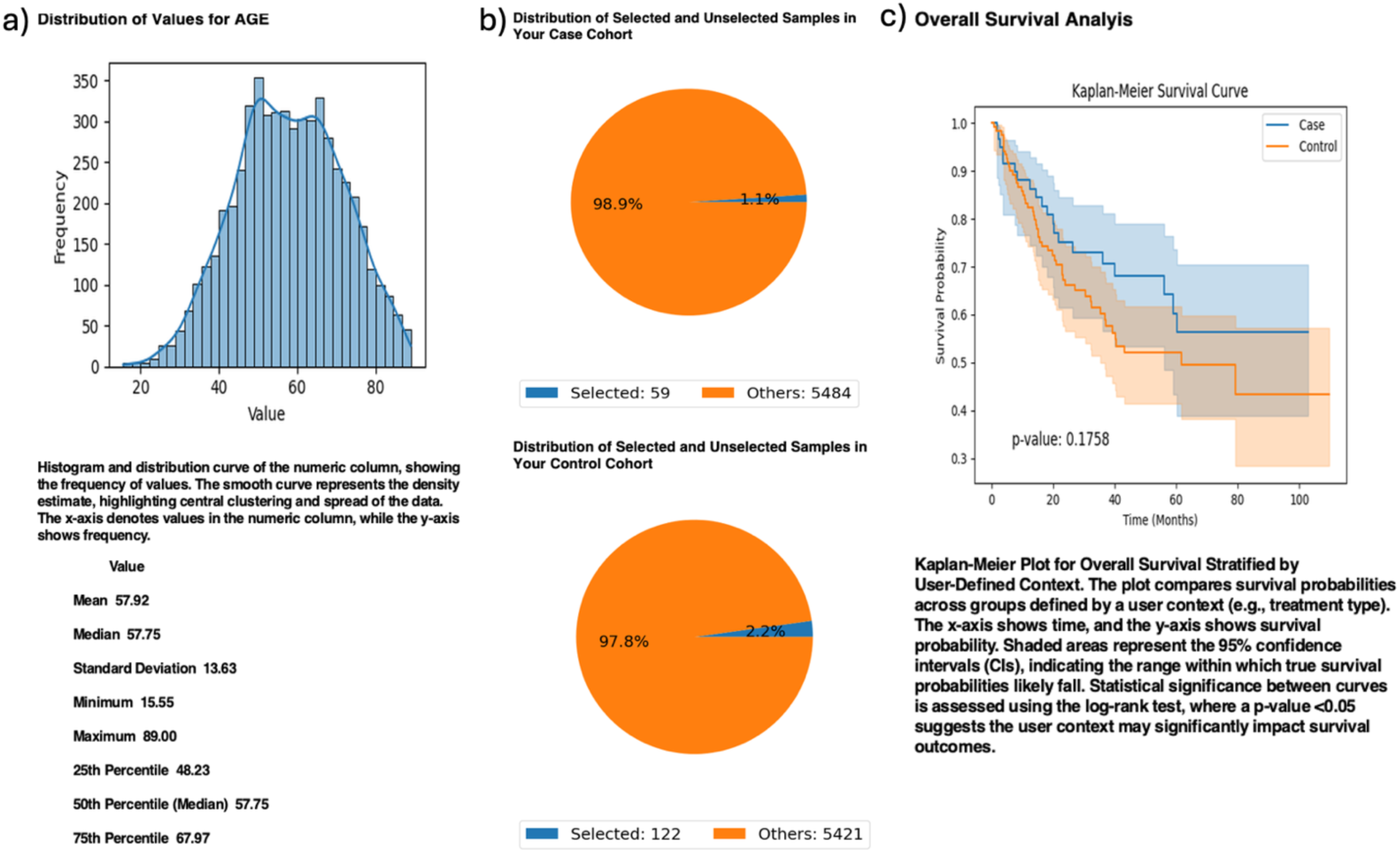
AI-HOPE-PI3K analysis of PTEN-mutated colorectal cancer (CRC) patients treated with FOLFOX chemotherapy, stratified by age group. This figure illustrates the application of AI-HOPE-PI3K to assess survival outcomes in PTEN-mutated CRC patients treated with the FOLFOX regimen (Fluorouracil, Leucovorin, Oxaliplatin), comparing early-onset (<50 years) and late-onset (>50 years) cases. a) A histogram depicts the distribution of patient age across the dataset, with a mean of 57.92 years and a median of 57.75. The smooth curve overlay shows central clustering and spread of the data, contextualizing the age cutoff used for cohort stratification. b) Pie charts illustrate the cohort selection process following natural language query execution. The case cohort includes 59 early-onset CRC patients with PTEN mutations treated with FOLFOX (1.1% of the dataset), while the control cohort includes 122 late-onset CRC patients with the same molecular and treatment profile (2.2%). c) Kaplan-Meier survival analysis compares overall survival between early-onset and late-onset groups. Although early-onset patients demonstrated a trend toward improved survival, the difference was not statistically significant (p = 0.1758). Shaded regions represent the 95% confidence intervals, suggesting overlapping survival distributions and no conclusive evidence of age-based survival differences in this PTEN-mutated, FOLFOX-treated CRC subgroup.

**Figure S5.**
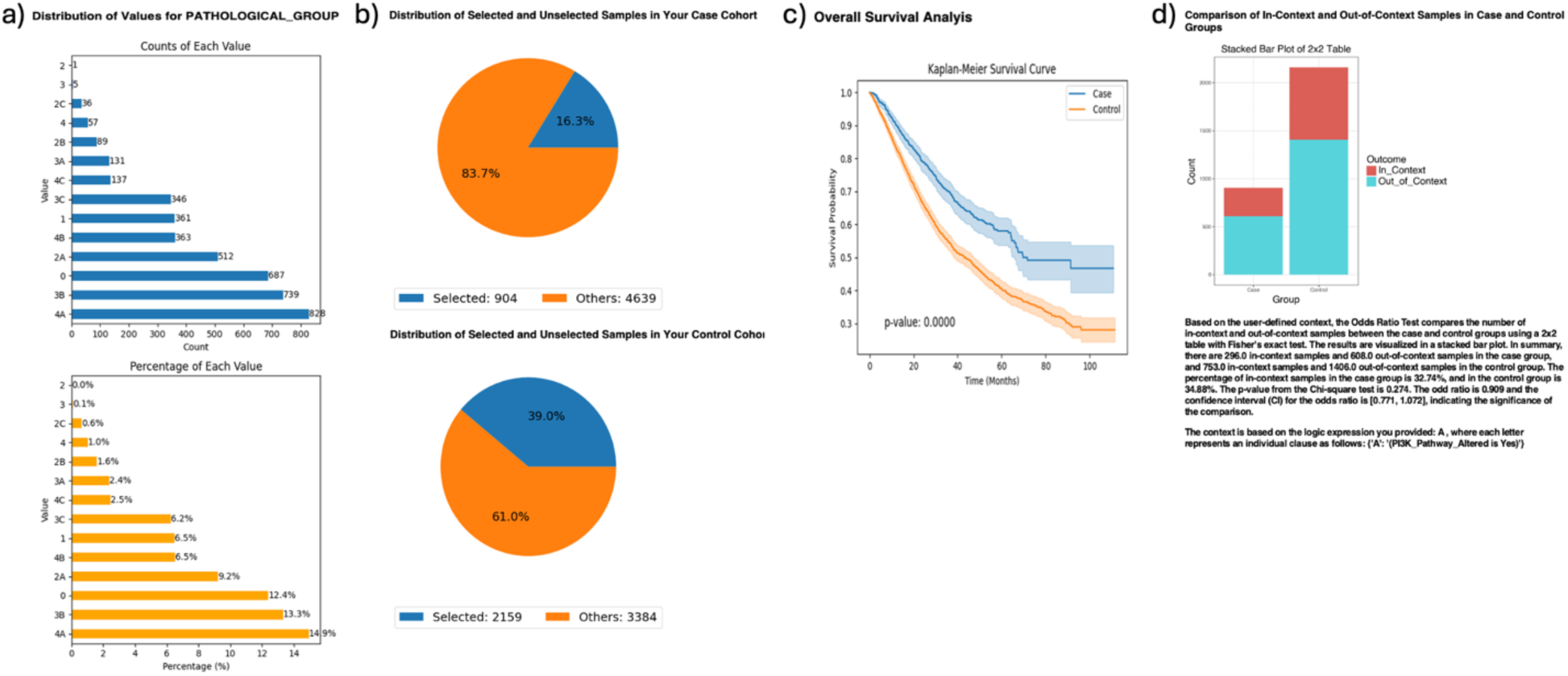
AI-HOPE-PI3K analysis of PI3K-altered colorectal cancer (CRC) stratified by pathological stage group: Early (Stage 0–2C) vs. Advanced (Stage 3– 4C) disease. This figure demonstrates the application of AI-HOPE-PI3K to evaluate survival outcomes and molecular enrichment among CRC patients harboring PI3K pathway alterations, stratified by pathological stage at diagnosis. All patients received FOLFOX chemotherapy (Fluorouracil, Leucovorin, Oxaliplatin). a) Bar plots depict the distribution of pathological stage groups across the dataset. Early-stage cases (Stage 0, 1, 2, 2A, 2B, 2C; labeled X0) represent the majority of samples (54.2%), while advanced-stage cases (Stage 3A–4C; labeled X1) account for 45.8%. The upper panel shows the sample count per stage category, and the lower panel presents their proportional representation. b) Pie charts visualize the cohort selection process following natural language–driven filtering. The case cohort includes 628 early-stage CRC patients with PI3K pathway alterations treated with FOLFOX (11.3% of the dataset), while the control cohort consists of 507 advanced-stage PI3K-altered CRC patients treated with the same regimen (9.1%). These distributions contextualize cohort sizes and selection logic. c) Kaplan-Meier survival curves compare overall survival between the two groups. Despite a visual trend suggesting improved survival in the early-stage PI3K-altered group, the difference was not statistically significant (p = 0.1267). Shaded regions represent 95% confidence intervals. d) A multivariate odds ratio analysis was conducted to assess the association between PI3K pathway alterations and pathological stage.

